# Personalised structural connectomics for moderate-to-severe traumatic brain injury

**DOI:** 10.1101/2022.03.02.22271654

**Authors:** Phoebe Imms, Adam Clemente, Evelyn Deutscher, Ahmed M. Radwan, Hamed Akhlaghi, Paul Beech, Peter H Wilson, Andrei Irimia, Govinda Poudel, Juan F Domínguez D, Karen Caeyenberghs

## Abstract

Graph theoretical analysis of the structural connectome has been employed successfully to characterise brain network alterations in patients with traumatic brain injury (TBI). However, heterogeneity in neuropathology is a well-known issue in the TBI population, such that group comparisons of patients against controls are confounded by within-group variability. Recently, novel single-subject profiling approaches have been developed to capture inter-patient heterogeneity. We present a personalised connectomics approach that examines structural brain alterations in six chronic patients with moderate-to-severe TBI who underwent anatomical and diffusion magnetic resonance imaging (MRI). We generated individualised profiles of lesion characteristics and network measures (including personalised graph metric ‘GraphMe’ plots, and nodal and edge-based brain network alterations) and compared them against healthy reference cases (N=12) to assess brain damage qualitatively and quantitatively at the individual level. Our findings revealed clinically significant alterations of brain networks with high variability between patients. Our profiling can be used by clinicians to formulate a neuroscience-guided integrative rehabilitation program for TBI patients, and for designing personalised rehabilitation protocols based on their unique lesion load and connectome.

---

Moderate-to-severe traumatic brain injury (TBI) can result in diverse focal lesions and white matter pathology. The locations of these lesions greatly contribute to functional outcomes following TBI, whereby cognitive functions that rely on broadly distributed circuits in the brain are affected due to disruptions to axonal pathways and cortical structures^1,2,3^. In TBI patients, diffusion weighted MRI (dMRI) studies have shown altered topological properties of structural brain networks, as indexed by graph metrics at the group level^4-7^. In our recent meta-analysis^8^, we found that only two of 14 graph metrics (characteristic path length and normalised clustering coefficient) showed significant differences in TBI patients compared with controls, reflecting the heterogeneous nature of TBI patients. This heterogeneity, including complex structural profiles, variation in lesion location, severity, response to treatment, as well as varied secondary injury trajectories, poses a challenge for the prediction of functional and cognitive symptoms of TBI patients. As a result, there is growing impetus for subject-tailored approaches that enable injury characterization and treatment planning^9-11^.

Recent studies have addressed heterogeneity in clinical cohorts by performing individualised analyses of dMRI-derived fractional anisotropy (FA), T1-derived cortical thickness, and streamline counts^11-13^ at the level of white matter (WM) tracts or grey matter (GM) regions, respectively. For example, Lv and colleagues^13^ found no group consensus in anatomic locations of lower FA and reduced cortical thickness in schizophrenia patients, and as such group-level FA and cortical thickness maps were not representative of individuals. To date, however, few studies have analysed brain networks at the level of individual patients, an approach known as *personalised connectomics*^10^.

Pioneered by Irimia and colleagues^10^, personalised connectomics enables the use of an individual’s brain network as a ‘fingerprint’ of brain network topology^14,15^. Personalised connectomics allows the visualization of individual white matter atrophy profiles (as indexed by dMRI-inferred streamline counts) using circular plots and considering patients scores relative to a healthy cohort. These individualized graphs can be used by clinicians to develop personalised rehabilitation programs, by detailing network level abnormalities^9^ that may indicate specific cognitive deficits following injury. No study to date has examined TBI patients’ network alterations using graph metrics, whereby a literature-driven selection of graph metrics that summarise segregation, integration, and centrality are represented for individual patients^16^. Since graph metrics were recently shown to have prognostic potential^7,17^, this type of approach could provide valuable information to clinicians, leading to neuroimaging-guided strategies to improve functional outcomes of TBI patients. However, personalised connectomics in moderate to severe TBI cohorts with diverse brain injuries pose a serious technical challenge, as the available tools for MRI processing to generate connectomes fail in such^18^.

The present study introduces personalized measurement and analysis of individual connectomic profiles in five chronic moderate-to-severe TBI patients with varying lesion loads, mechanisms of injury, age at injury, and burden of neural/cognitive symptoms. Our implementation extends current methods by addressing the long-standing and prominent challenge of analysing TBI structural profiles when automatic sub/cortical segmentation or parcellation of MRIs fail in the presence of lesions^18^. Significantly, this problem is addressed here by synergizing connectomic analysis with *virtual brain repair*, where the lesion is replaced by healthy looking tissue in the T1-weighted images (lesion inpainting). The capabilities of our implementation of personalised connectomics in TBI include: (*i*) *lesion masking* undertaken in a semi-automated manner from anatomical T1 MRI scans to identify the affected brain regions in individual patients; (*ii*) the use of the recently developed *Virtual Brain Grafting* (VBG) toolbox to overcome the challenges of segmentation and parcellation of focal lesions using lesion inpainting^19^; (*iii*) graphical representation of the structural connectome using innovative tools for graph metric profiling (GraphMe plots) to delineate subject-specific changes in brain network integration, segregation, and centrality; and i*v*) regional assessment of network hub regions and edge alterations in individual TBI cases. Together, these innovative solutions overcome major, longstanding methodological impediments in the field of macroscale TBI profiling. Our implementation is the first to allow the comprehensive generation of lesion-aware connectomic profiles that may be used to inform clinical decision making, thus moving closer to the crucial aim of translating research findings into clinical practice.

## Materials and Methods

### Participants

Patients with chronic moderate-to-severe TBI were recruited from St Vincent’s Hospital in Melbourne (SVHM). The definition of moderate-to-severe TBI was based on (i) a Glasgow Coma Scale score at the time of hospital admission between 3-12^20^; (ii) loss of consciousness of at least 30 minutes; (iii) post-traumatic amnesia of at least 24 hours^21^: and (iv) positive findings of gross injury on MRIs as per evaluation by a neuroradiologist (PB). Patients who met the following inclusion criteria were contacted to take part in the study: (a) between 18 and 65 years of age; (b) no history of head injury prior to the TBI for which they were included in this study; (c) fluency in English, (d) no history of psychiatric illness prior to the TBI, and (e) no contra-indications for MRI. Ten moderate to severe TBI patients who had sustained closed head injuries due to sports or motor-vehicle accidents more than 6 months prior to the study were recruited. Informed written consent was obtained from each subject in accordance with the Declaration of Helsinki. Due to time constraints during scanning, dMRI were not acquired from four TBI patients, who were subsequently removed from further analysis (see Table 1). One further participant was removed from personalised connectome construction due to excess movement in the scanner during dMRI, which caused a severe motion artifact (see Supplementary Material 2). For the reference group, 12 healthy controls were recruited from the general population using flyers and the snowball method (see Table 1). Ethical approval was granted by the SVHM ethics committee for human research (project #250/17).

**Table 1.** Participant Demographics and Injury Characteristics

| ID | Age<sup>1</sup> | Sex | TSI<sup>2</sup> | Mechanism | Pathology (at time of study)<sup>3</sup> | DAI<sup>4</sup> |
| --- | --- | --- | --- | --- | --- | --- |
| HC | 35.7<br>±<br>11.4 | M = 4<br>F = 8 | - | - | No incidental or age-related findings, other than small deep white matter T2 hyperintensities (within normal limits for age). | - |
| TBI1 | 40's | M | 21y | Vehicle accident | Modest encephalomalacia in the (R) precentral gyrus | 0 |
| TBI2 | 40's | M | 15y | Vehicle accident | Severe encephalomalacia involving both ant. F and inf. F lobes, (R) T lobe and (R) parietotemporal region extending to the (R) post. F lobe. Focal T1 hypointensities in the anteromedial portion of the (L) thalamus. Encephalomalacia and T1 hypointensity on the ant. body and genu of the corpus callosum. | 2 |
| TBI3 | 40's | F | 3y | Fall | Bilateral ant. and inf. F encephalomalacia, (R) greater than (L), and (R) ant. T encephalomalacia. Small deep white matter T2 hyperintensities med. (R) P lobe, likely associated with non-haemorrhagic oedema. Small focal T1 hypointensity in the ant. body of the corpus callosum. | 2 |
| TBI4 | 30's | F | 15y | Fall | Bilateral inf. F and (L) ant. T encephalomalacia. Modest encephalomalacia in the (L) sup. F gyrus. (R) F ventriculostomy with underlying ventricular drain tract. | 0/1 |
| TBI5 | 50's | M | 18y | Vehicle accident | Two small (<2mm<sup>3</sup>) deep white matter T2 hyperintensities in the (R) P lobe (within normal limits for age). | 0 |
| TBI6 | 30's | F | 5y | Fall | Small T1 hypointensity in the splenium of corpus callosum. Approx. 6 scattered punctate T2 hyperintensities in both cerebral hemispheres. | 2 |

<sup>1</sup>Age: Shown in 10-year age bracket to minimise identifiable information, HC age is in mean ± standard deviation. <sup>2</sup>TSI: Time since injury; <sup>3</sup>Abbreviations: (R) = right, (L) = left, ant. = anterior, post. = posterior, inf. = inferior, mid. = middle, med. = medial, sup. = superior, F = frontal, P = parietal, O =
occipital, T = temporal. <sup>4</sup>Grading of diffuse axonal injury (DAI) occurred according to Adams and colleagues<sup>22</sup>; a grade of ‘0’ indicates no confirmed DAI present, ‘1’ indicates DAI present in white matter of cerebral hemispheres, corpus callosum, brainstem, cerebellum; ‘2’ indicates there is also a focal lesion in corpus callosum; and ‘3’ identifies an additional lesion in dorsolateral quadrants of brainstem.

### Data acquisition

MRI scans were acquired at the Royal Children’s Hospital using a 3T Siemens PRISMA with a 64-channel head coil. dMRI data were acquired using a single-shot echo planar imaging sequence (twice-reinforced spin echo, multi-band acceleration factor of 2, 70 contiguous sagittal slices) and a high angular resolution diffusion imaging (HARDI) gradient scheme with 66 non-collinear gradient directions (total acquisition time (TA) = 6:17 mins, *b* = 3000 s/mm^2^, field of view (FOV) = 260 mm^2^, voxel size = 2.3 mm isotropic, repetition time (TR) = 3500 ms, echo time (TE) = 67 ms, seven volumes with *b* = 0, two reverse phase-encoded volumes with *b* = 0, *b* being the constant of diffusion weighting). T1-weighted MRIs were also acquired using a magnetisation-prepared rapid acquisition gradient-echo (TA = 5:48 mins, 208 contiguous slices, FOV = 256 mm^2^, voxel size = 0.8 mm isotropic, TR = 2100 ms, TE = 2.22 ms, flip angle = 8°).

### Lesion masking

Manual lesion delineation for computation of lesion load and for improvement of anatomical segmentation was performed by an assessor (ED), who was trained in lesion identification by neuroradiologist (PB). Lesions were drawn in the T1 native space using Fsleyes version 0.27.3 in FSL version 6.0.1 (https://fsl.fmrib.ox.ac.uk/fsl/fslwiki). An in-house systematic search method and lesion identification protocol was implemented by JD, KC, ED, and PB. Abnormalities resulting in tissue loss, such as regions of encephalomalacia and damage from surgical drainage tracts were included in binarised lesion masks. Enlarged ventricles and hyperintensities often occurring in proximity to the skull (e.g., from surgical craniotomies) were not included in the lesion masks. Lesion load was computed (in cm^3^) as the total volume of the binary lesion masks in FSL.

### Personalised connectome construction

Our connectome processing pipeline is showcased in Figure 1, Supplementary Material 1, and in our previous publication^23^. Our personalised connectomics implementation performs state-of-the-art, single-subject analyses of structural MRI scans. Briefly, raw dMRI data were processed using MRtrix3Tissue (v5.2.8; https://3tissue.github.io), a fork of MRtrix3^24^. White matter fibre orientation distributions were estimated using single-shell 3-tissue constrained spherical deconvolution (SS3T-CSD)^25,26^. Whole-brain, anatomically-constrained tractography was performed^27^ and twenty-two million streamlines were generated per subject^28^. The SIFT2 algorithm was applied to match the fibre density of the reconstructed streamlines to that of the underlying white matter structures^28-30^. Thus, edges encode filtered streamlines count.

**Fig. 1.**
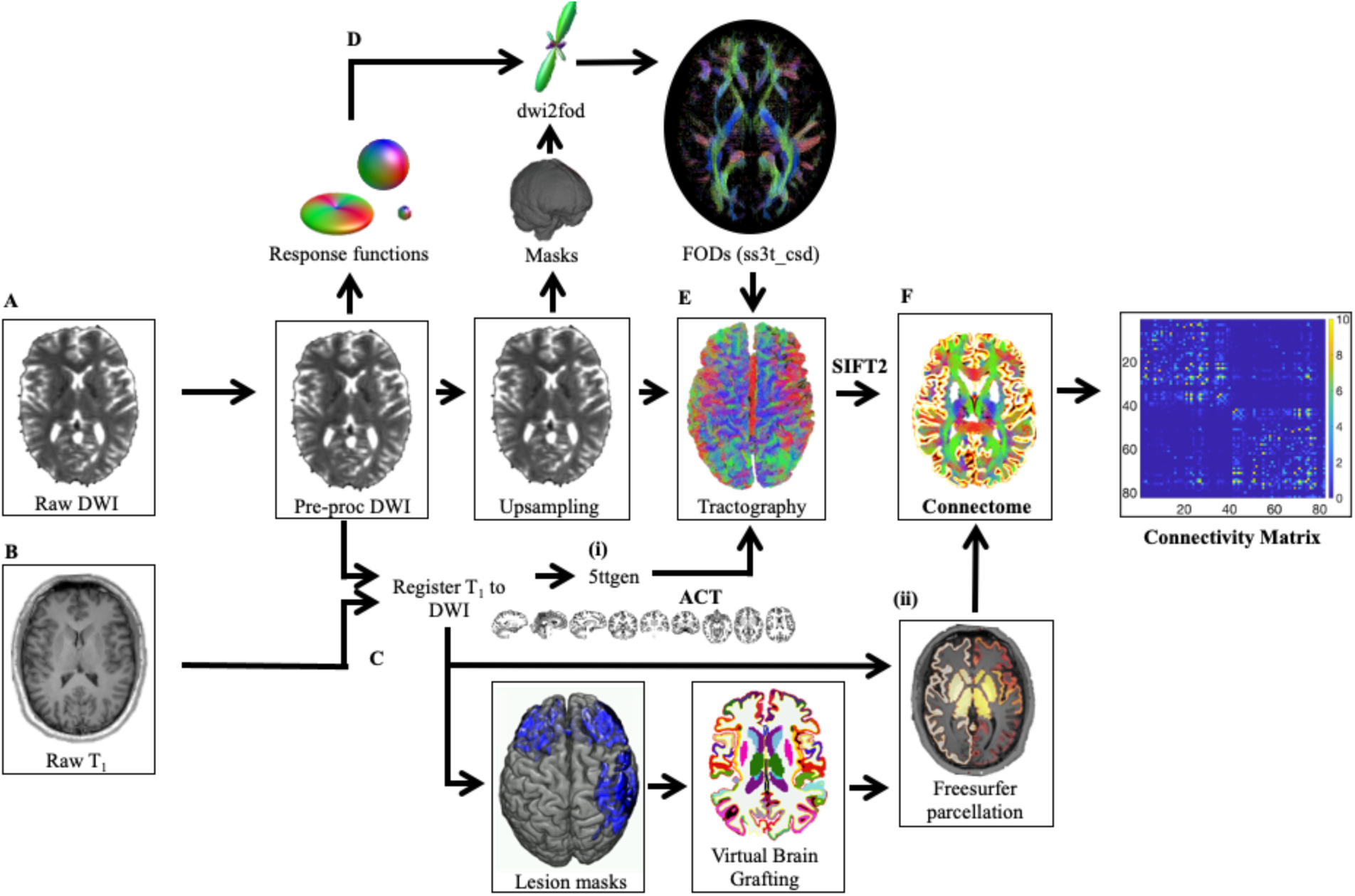
Overview of the processing pipeline for connectome mapping. (A) In the raw diffusion images, noise^33,34^, Gibbs ringing artefacts^35^, as well as distortions induced by motion, eddy current artefacts and EPI/susceptibility distortions were detected and corrected^36,37^. (B) Concurrently, T1 volumes were registered to diffusion volumes. The advanced normalisation tools package (ANTS)^38^ was used to remove non-brain structures from the T1 weighted images for white matter extraction^39^. FSL FLIRT^40,41^ was used to perform the boundary-based registration between brain-extracted anatomical and diffusion images. Registered images are provided to (i) 5ttgen (brain extracted), to create priors for anatomically constrained tractography (ACT), and (ii) FreeSurfer (non-brain extracted), to parcellate the nodes for the connectome analysis. All subcortical grey-matter structures were segmented^42^; image intensity normalised^43^; pial surfaces and the grey-white matter boundaries estimated^44^; and the entire brain “inflated” to smooth the gyri and sulci^45^. (C) Lesion maps of subjects who failed the quality assessment after FreeSurfer parcellation were provided along with the T1 image to VBG. (D) Average response functions for white matter, grey matter, and cerebrospinal fluid were estimated from the dMRI data using an automated unsupervised approach^46,47^. Pre-processed data were upsampled to a voxel size of 1.3 mm^3^ to assume higher spatial resolution for image registration before binary masks were created. Fibre orientation distributions (FODs) were estimated from the group average response functions on upsampled images, and corrected for intensity inhomogeneity and global intensity level differences^48^. (E) Anatomically constrained tractography (ACT) was performed using the FODs from (D) and the 5ttgen images from (B(i)). The FOD cut-off threshold, step size, and angle were determined to attain a reasonable trade-off between false negatives and false positives (seed points=dynamic; maximum length=250mm; minimum length=5 mm; step size=1.25; angle=45°; FOD amplitude = 0.08). Spherically informed filtering of tractograms (SIFT2) is applied to make the weight of the streamlines proportional to the underlying fibre orientation distribution. (F) The connectome is created using the FreeSurfer parcellation and the sifted tractogram.

T1 anatomical MRIs were parcellated into 84 regions of the Desikan-Killiney atlas^31^ using FreeSurfer’s *recon-all* function (v6.0; http://surfer.nmr.mgh.harvard.edu/)^32^. Two patients (TBI3 and TBI4) had significant segmentation failures due to gross pathology, and were therefore processed utilising VBG v0.37^19^. Rather than lesion masking and manual editing which are subjective and time-consuming, VBG automatically fills uni- and bilateral brain lesions using synthetic healthy donor tissue to permit or to improve segmentation. To illustrate the performance of VBG in TBI, we included a report on VBG outcome for patient TBI2, who was excluded from personalised connectomics due to movement during HARDI acquisition but otherwise had a quality control compliant T1-weighted volume (Supplementary Material 3). Given VBG artificially reconstructs lesioned nodes, part of our quality control also included ensuring streamlines were not aberrantly assigned to these nodes. Connectivity matrices were generated using edge weights from SIFT2 and nodes defined as brain regions from FreeSurfer and VBG. Global network properties were quantified in terms of *strength, global efficiency, characteristic path length, navigation efficiency, average local efficiency, clustering coefficient, normalised clustering coefficient*, and *average betweenness centrality* (Table 2) using the Brain Connectivity Toolbox^16^. These graph metrics were chosen from all available metrics as the most clinically informative/intuitive according to previous findings from our meta-analysis^8^. Grading of diffuse axonal injury (DAI) was performed by expert raters PB and ED (Table 1)^22^.

**Table 2.** Graph Metric Descriptions and Interpretations

| Graph Metric | Description | Higher values mean... | Previous studies<br>(Adult msTBI#) |
| --- | --- | --- | --- |
| *Integration* |  |  |  |
| *Characteristic Path Length* | The shortest path is the fastest and most direct communication pathway between two network nodes. Characteristic path length is defined as the average shortest path length between all node pairs in a network<sup>49</sup>. | A higher characteristic path length indicates that the fastest communication pathways between regions are, on average, longer and **less** efficient. | **Higher** CPL<sup>4,5,50</sup> |
| *Global Efficiency* | The inverse average shortest path efficiency between all possible pairs of nodes in a network, where efficiency is computed as the inverse of shortest the path length<sup>51</sup>. | A higher global efficiency indicates a **greater** capacity for efficient integration of information (in parallel) across the network. | **Lower** global efficiency<sup>4</sup> |
| *Navigation Efficiency* | Navigation paths use a decentralised and geometrically greedy heuristic<sup>52</sup>. Navigation efficiency is defined as the average navigation path efficiency between all possible pairs of nodes in a network<sup>53</sup>. | Higher navigation efficiency indicates **greater** capacity for efficient integration of information across the network. | Not yet investigated, but **lower** navigation efficiency observed in stroke patients<sup>54</sup> |
| *Segregation* |  |  |  |
| *Clustering Coefficient* | The number of existing connections between the neighbours of a node, divided by all the possible connections, calculated for each node individually and averaged across the entire network<sup>49</sup>. | A higher average clustering coefficient implies that a greater proportion of connections are made between nodes neighbours, compared to the connections possible, and indicates more clustered connectivity around individual nodes. | **Lower** clustering coefficient<sup>6,50</sup> |
| *Normalised Clustering Coefficient* | Clustering coefficient of the network normalised to a random network. | Higher normalised clustering indicates **higher** local specialization, with a value of 1 being equivalent to a random network. If greater than 1, the network has greater than random clustering. There may be a point of diminishing returns, where greater local specialization comes at the **cost** of communication efficiency. | **Higher** normalised clustering*<sup>55,56</sup> |
| *Local Efficiency* | The local efficiency is the average of inverse shortest path length in a local area. Mean local efficiency is the efficiency of each node in the network averaged over the total number of nodes<sup>51</sup>. | A higher local efficiency means **greater** capacity for integration between the immediate neighbours of a given node. | **Higher** local efficiency<sup>57</sup>; **AND/OR lower** local efficiency<sup>58</sup> |
| *Centrality* |  |  |  |

| Graph Metric | Description | Higher values mean... | Previous studies<br>(*Adult msTBI*<sup>#</sup>) |
| --- | --- | --- | --- |
| *Strength* | The strength of a node is the sum of the weights of its edges. Mean strength is the average of all the normalised strength values across each node of the network. | A higher strength indicates a **greater** average edge weight for each node. | **Lower** strength<sup>6</sup> |
| *Betweenness Centrality* | The proportion of shortest paths that pass-through node i between its neighboring nodes, calculated for each node and averaged across the network<sup>59</sup>. | Higher betweenness centrality means that node lies on more shortest paths in the network pass through it, and this that node is **more central** and important to the network. A high network/average betweenness centrality indicates a high number of nodes that are central to shortest paths. | **Higher** betweenness centrality<sup>55</sup> * |

<sup>#</sup>msTBI: moderate-severe traumatic brain injury. \* Note: this study is of young adults and children with TBI – no adult TBI study found significant alterations or examined this metric.

### Brain network profiles

Graph metrics spiderplots (‘GraphMe’ plots) show results for each TBI patient in a concise and intuitive manner relative to mean scores from the healthy controls with 95% confidence intervals. Selected graph metrics (characteristic path length, normalised clustering coefficient, and betweenness centrality – Table 2) were inverted (1/*x*) to facilitate interpretation (so that *higher* scores on any graph metric denote *better* brain network structure). Graph metrics of individual patients were categorised as follows: *normal* (if the scores/metrics fell within the 95% confidence interval); *supra-normal* (higher than the 95% confidence interval); or *infra-normal* (lower than the 95% confidence interval)^13^.

### Regional brain network analyses

A key component of personalised connectomics is to localise network alterations in the brain relative to a healthy cohort. Nodal hubs and weakest edges were also examined for each individual patient based on comparison to the healthy controls. *Betweenness centrality* was used to identify brain regions essential for communication within the brain network^16,59^. The top 10% (*n*=8) highest-scoring nodes were identified as hubs; for the healthy control group these are shown in Figure 2, and for the TBI patients these are shown in Figures 3(a) to (d).

**Fig. 2.**
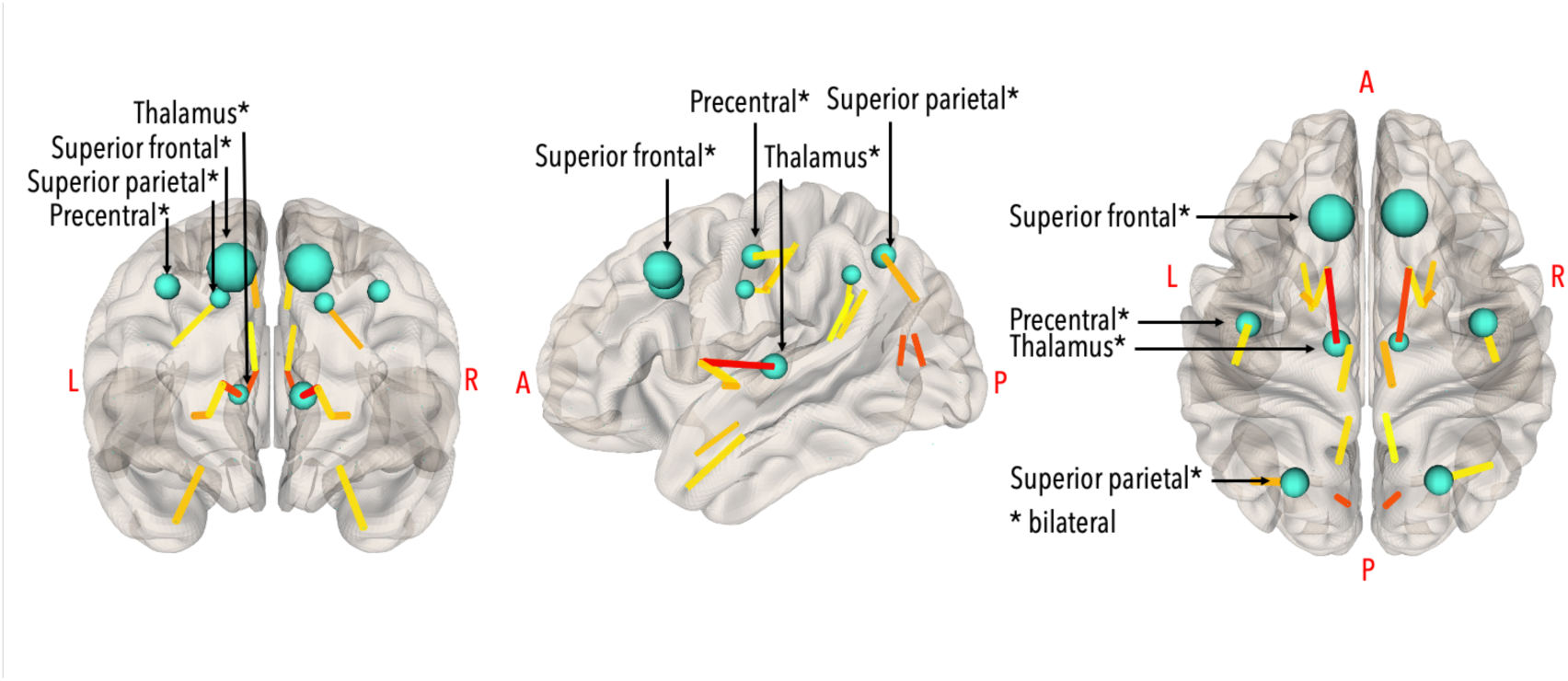
Healthy control hubs (top 10% of nodes with highest betweenness centrality), in teal. Larger nodes represent higher values. Hubs (bilaterally) were the superior frontal gyrus (BC_left_=1493; BC_right_=1533), superior parietal gyrus (BC_left_=610; BC_right_=665), precentral gyrus (BC_left_=588; BC_right_=616), and thalamus (BC_left_=336; BC_right_=346). The strongest edges (0.5^th^ percentile) are coloured by strength (yellow=weaker; red=stronger). Visualisation in NeuroMArVL (https://immersive.erc.monash.edu/neuromarvl/).

**Fig. 3.**
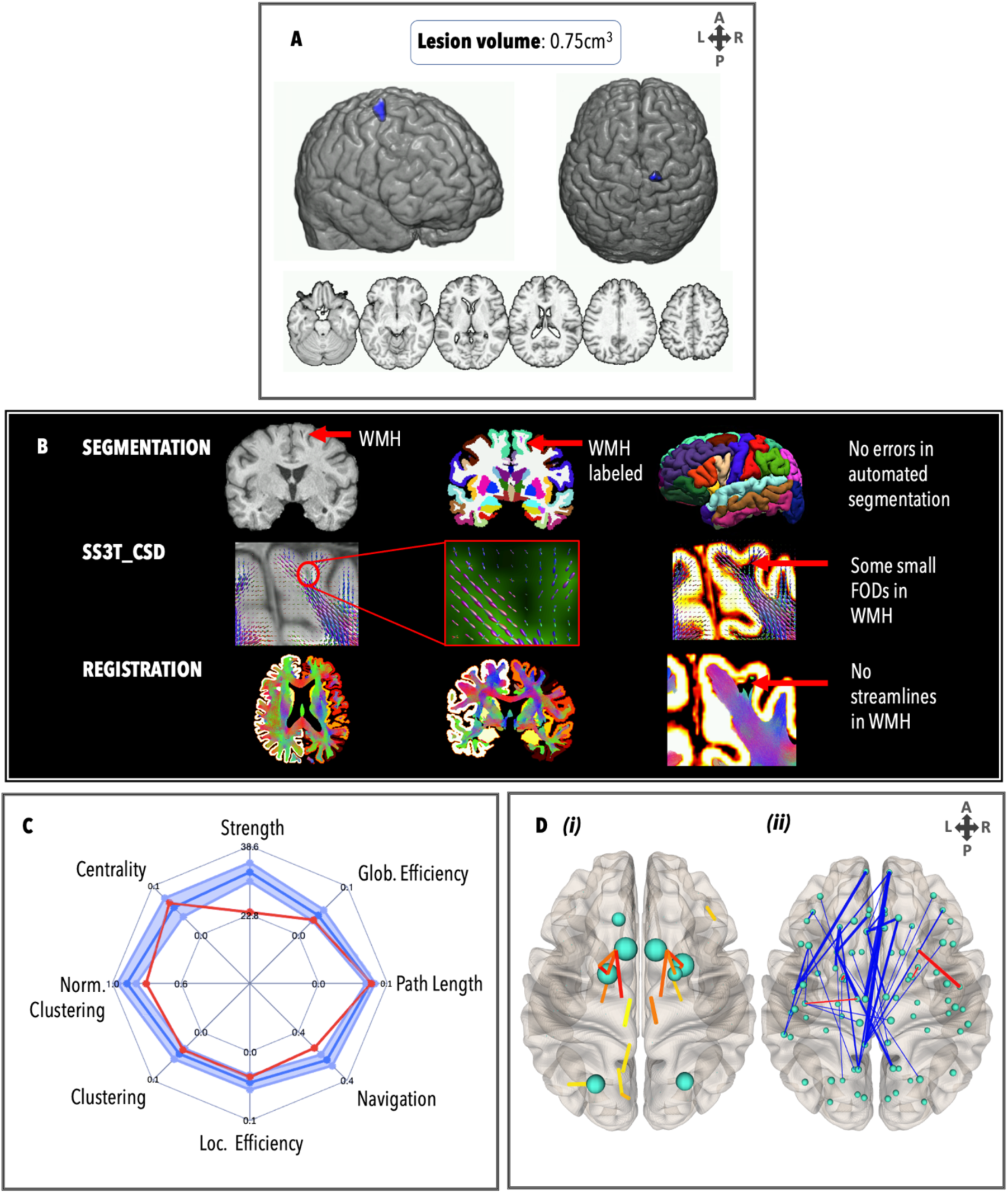
Personalised connectome profile for TBI1 including (A) lesion profile; (B) quality assessment; (C) radar plot showing the patient’s personalised connectome profile (red indicates patient’s scores, dark blue indicates healthy control average and the 95% Cis are represented by the blue shade); and (D) regional analysis (blue: edges lower than the healthy control average; red: edges stronger than the healthy control average; thicker edges: more standard deviations away from the healthy mean).

### Z-score Matrix for Edge Analysis

An edge analysis scrutinised the white matter connections that drive overall differences in the network properties in greater detail^60^. A *z*-score matrix *Z*_*i,j*_ was derived, which describes the distance from the healthy control mean, divided by the healthy control standard deviation, between each subject’s connectivity matrix *T*_*i,j*_ and the controls *H*_*i,j*_ according to equations from a previous edgewise analysis^60^:

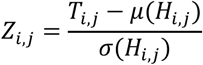

Positive scores represent stronger edges in the TBI patient compared to controls, while negative scores represent weaker edges. To visualise the *z*-score matrix, only edges with a score >4 (i.e., edges with weights that are 4 standard deviations away from the healthy control mean, corresponding to a highly conservative *p*<.0001) are displayed on a glass brain, while all other edges are discarded.

## Results

### TBI1

TBI1 (Figure 3) had a relatively small lesion load (0.75 cm^3^) spanning the posterior segment of the right superior frontal gyrus and right precentral gyrus, and a DAI grade of 0. Registration between structural and diffusion images was unaffected by this lesion. There were no failures in the FreeSurfer pipeline and there was no need for VBG. FODs were generated at the site of the lesion (see red arrow) but did not meet streamline criteria for ACT. The GraphMe plot indicated that TBI1 has slightly weaker integration than healthy controls, including infra-normal lower navigation efficiency, as well as lower strength and clustering coefficient. Regional brain network analyses revealed, interestingly, that the right superior frontal gyrus was not a hub node in TBI1. In addition, four other alterations in the hub arrangement for TBI1 were observed, whereby the nucleus accumbens (BC_left_=1570; BC_right_=1546), palladium (BC_left_=1382; BC_right_=978) and right putamen were bilateral hubs (BC_right_=1578), and the bilateral precentral gyri, thalami, and right superior frontal gyrus did not meet the hub threshold. Weaker edges (*z* < 4) (*n*=43 out of a total of 7056 edges) were observed projecting across frontal, parietal, temporal, and subcortical areas, in particular the edges between (A) the left posterior cingulate cortex and the right frontal pole (*z*=-8.32); (B) the left superior temporal gyrus and the left frontal pole (*z*=-6.66); (C) the left lateral orbitofrontal gyrus and the left temporal pole (*z*=-7.98); and (D) the left medial frontal and left temporal pole (*z*=-6.90). Some stronger (*z* > 4) edges (*n*=4) were also observed, including the connection between the right superior temporal gyrus and the right temporal pole (*z*=5.92).

### TBI3

TBI3 (Figure 4) had a relatively large lesion load (15.46cm^3^) involving primarily frontal regions (predominantly on the left), white matter hyperintensities in the medial right parietal lobe and the corpus callosum, and a DAI grade of 2. Prior to VBG, 10 nodes failed the quality assessment: VBG repaired 9 nodes for parcellation. Registration showed that streamlines were not assigned to lesioned nodes. The GraphMe plot demonstrated an infra-normal graph metric profile in all domains except centrality, which was normal. Two hub alterations were observed, whereby the bilateral putamen (BC_left_=871; BC_right_=932) were hubs, and the bilateral precentral gyri were not. Weaker edges (*n*=62) projected across the whole brain, especially the frontal regions, including between the left frontal pole and the left middle temporal gyrus (*z*=8.45), right superior frontal gyrus (*z*=8.12), right lateral orbitofrontal gyrus (*z*=8.17), and right putamen (*z*=8.41); between left medial orbitofrontal gyrus and the left amygdala (*z*=8.67); right lateral orbitofrontal gyrus and right nucleus accumbens (*z*=8.03) and right caudate nucleus (*z*=8.33); and right pars orbitalis and right lingual gyrus (*z*=8.10). No stronger edges were observed.

**Fig. 4.**
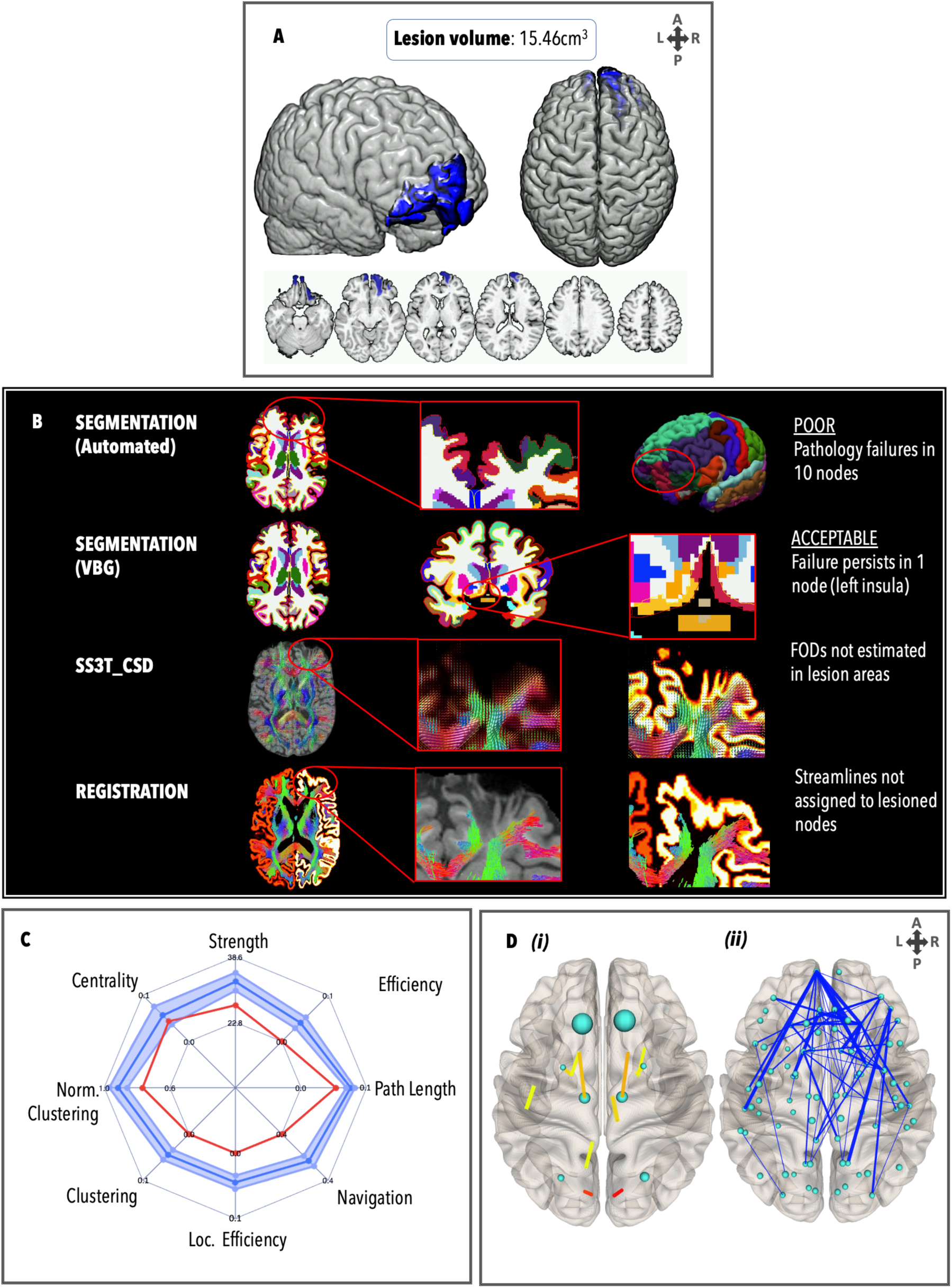
Personalised connectome profile for TBI3 including (A) lesion profile, (B) quality assessment, (C) radar plot showing the patient’s personalised connectome profile, and (D) regional analysis (blue: edges weaker than healthy control average; thicker edges: more standard deviations away from the healthy mean).

### TBI4

TBI4 (Figure 5) had a relatively large lesion load (17.59 cm^3^) involving bilateral frontal lesions and right temporal lesions, white matter hyperintensities in the medial right parietal lobe and the corpus callosum. However, the DAI grade was low (0/1). Prior to VBG, 9 nodes failed the quality assessment. All lesions overlapping with these nodes were repaired by VBG. Alignment between VBG-repaired nodes and streamlines indicated that any aberrant streamlines generated in areas with oedema/haemorrhage were not assigned to lesioned nodes. This patient exhibited supranormal graph metrics in all domains except the normalised clustering coefficient of the network (which was normal), and centrality (which was infra-normal). Four alterations in the hub arrangement were observed, whereby the bilateral putamen (BC_left_=2246; BC_right_=1550), left palladium (BC_left_=1210) and left inferior parietal (BC_right_=902) were hubs, and the bilateral precentral gyri and thalamic regions were not hubs. Weaker edges (*n*=26) projected across the left hemisphere, including between the entorhinal and lingual gyrus (*z*=10.87), pericalcarine (*z*=9.55), superior parietal (*z*=9.46), and lateral occipital regions (*z*=9.33); the temporal pole and the insula (*z*=9.37); and the nucleus accumbens and the posterior cingulate cortex (*z*=6.71), insula (*z*=6.30), and rostral anterior cingulate cortex (*z*=5.98). Some stronger edges (*n*=4) were also observed in the right hemisphere, including between the pars triangularis and postcentral gyrus (*z*=5.92), the putamen and lateral orbitofrontal (*z*=4.86), the pallidum and the thalamus (*z*=4.24); and the left pallidum and the amygdala (*z*=4.55).

**Fig. 5.**
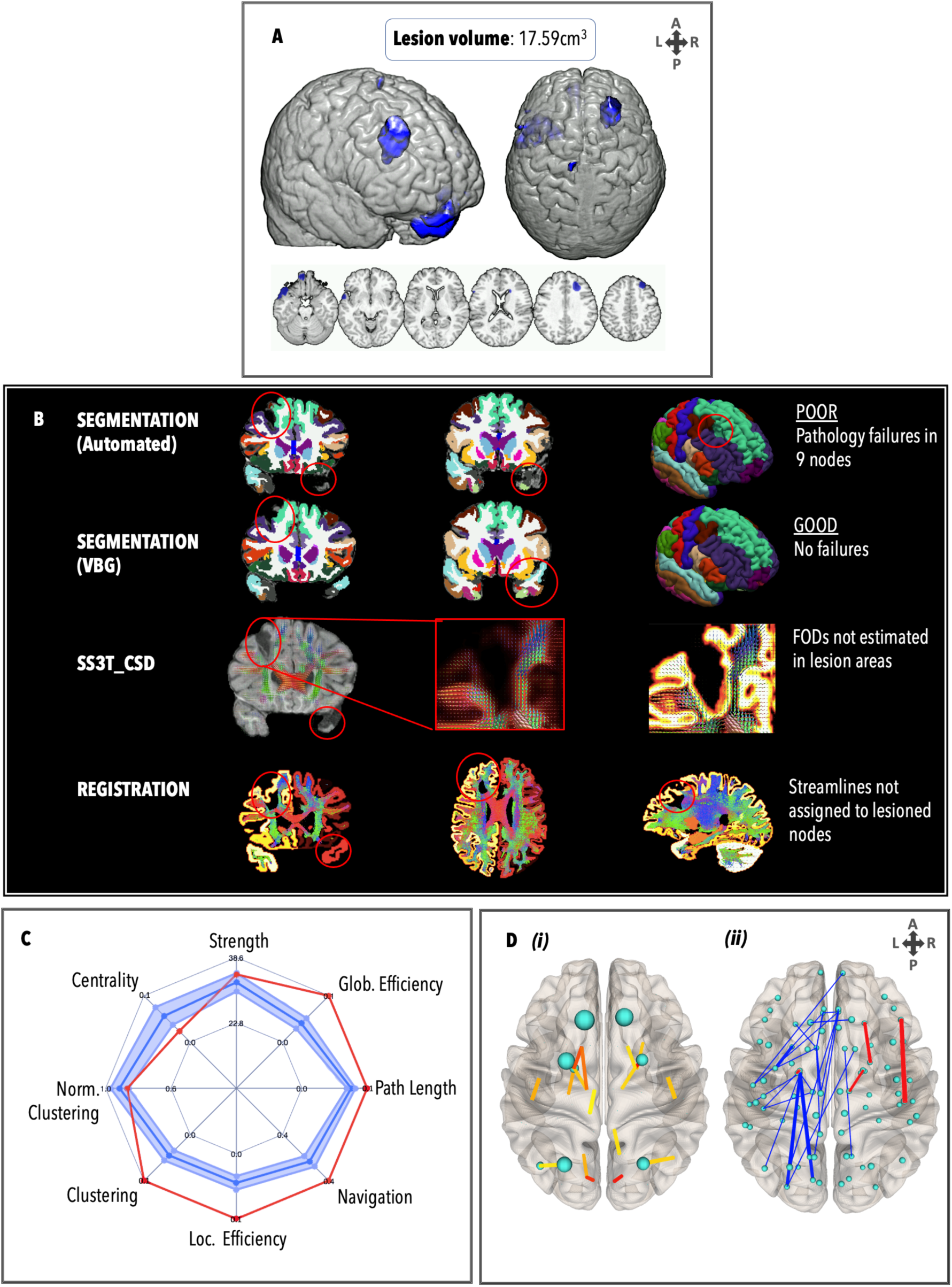
Personalised connectome profile for TBI5 including (A) lesion profile, (B) quality assessment, (C) radar plot showing the patient’s personalised connectome profile, and (D) regional analysis (blue: edges weaker than healthy control average; red: edges stronger than healthy control average; thicker edges: more standard deviations away from the healthy mean).

### TBI5

TBI5 (Figure 6) had no MRI-discernible lesion load and a DAI grade of 0. Many weaker edges were observed relative to healthy controls that connected the parietal, temporal, and subcortical lobes. There were no failures in the FreeSurfer pipeline, and no manual edits were necessary. FODs were generated correctly and registration between segmentation and tractography was free of error. The GraphMe plot revealed infra-normal strength, navigation, and normalised clustering. Two alterations in hub arrangement were observed, whereby the bilateral putamina were hubs (BC_left_=1182; BC_right_=1110), whereas the bilateral thalami were not. Weaker edges (*n*=33) projected inter-hemispherically across parietal, temporal, and subcortical areas. The weakest edges (compared to healthy controls) were in the left hemisphere, between the amygdala and the temporal pole (*z*=-7.19); the amygdala and the inferior temporal gyrus (*z*=-7.80); the inferior temporal gyrus and the hippocampus (*z*=-6.11); and the inferior temporal gyrus and the thalamus (*z*=-6.40). Edges stronger than in healthy controls (*n*=3) were also observed, including the set of connections between the left postcentral gyrus and the left lateral occipital gyrus (*z*=5.87).

**Fig. 6.**
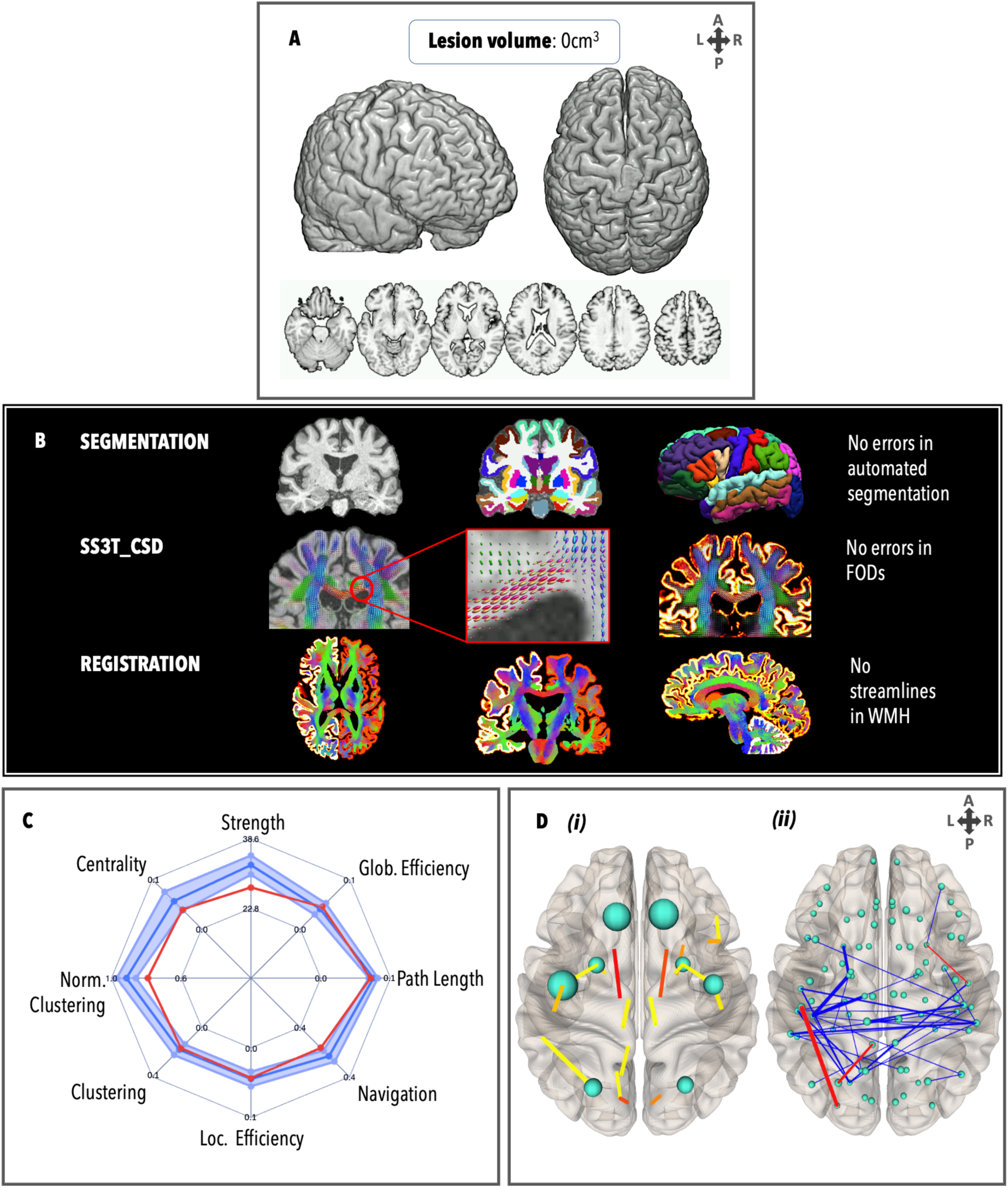
Personalised connectome profile for TBI5 including (A) lesion profile, (B) quality assessment, (C) radar plot showing the patient’s personalised connectome profile, and (D) regional analysis (blue=edges lower than healthy control average; red = edges stronger than healthy control average; and thicker edges=larger number of standard deviations away from healthy mean).

### TBI6

TBI6 (Figure 7) had a small lesion in the splenium of the corpus callosum (0.5 cm^3^), and a DAI grade of 2. There were no failures in the FreeSurfer pipeline, and no manual edits were necessary. FODs were generated at the site of the lesion but did not meet streamline criteria for ACT. The GraphMe plot showed that the global connectivity properties of TBI6 were normal. Three hub alterations were observed, whereby the right caudate nucleus (BC_right_=722), right hippocampus (BC_right_=606) and right inferior parietal gyrus (BC_right_=680) were hubs, and the bilateral precentral and right superior parietal regions were not hubs. No edges met the stringent threshold of being at least 4 standard deviations away from the healthy control mean.

**Fig. 7.**
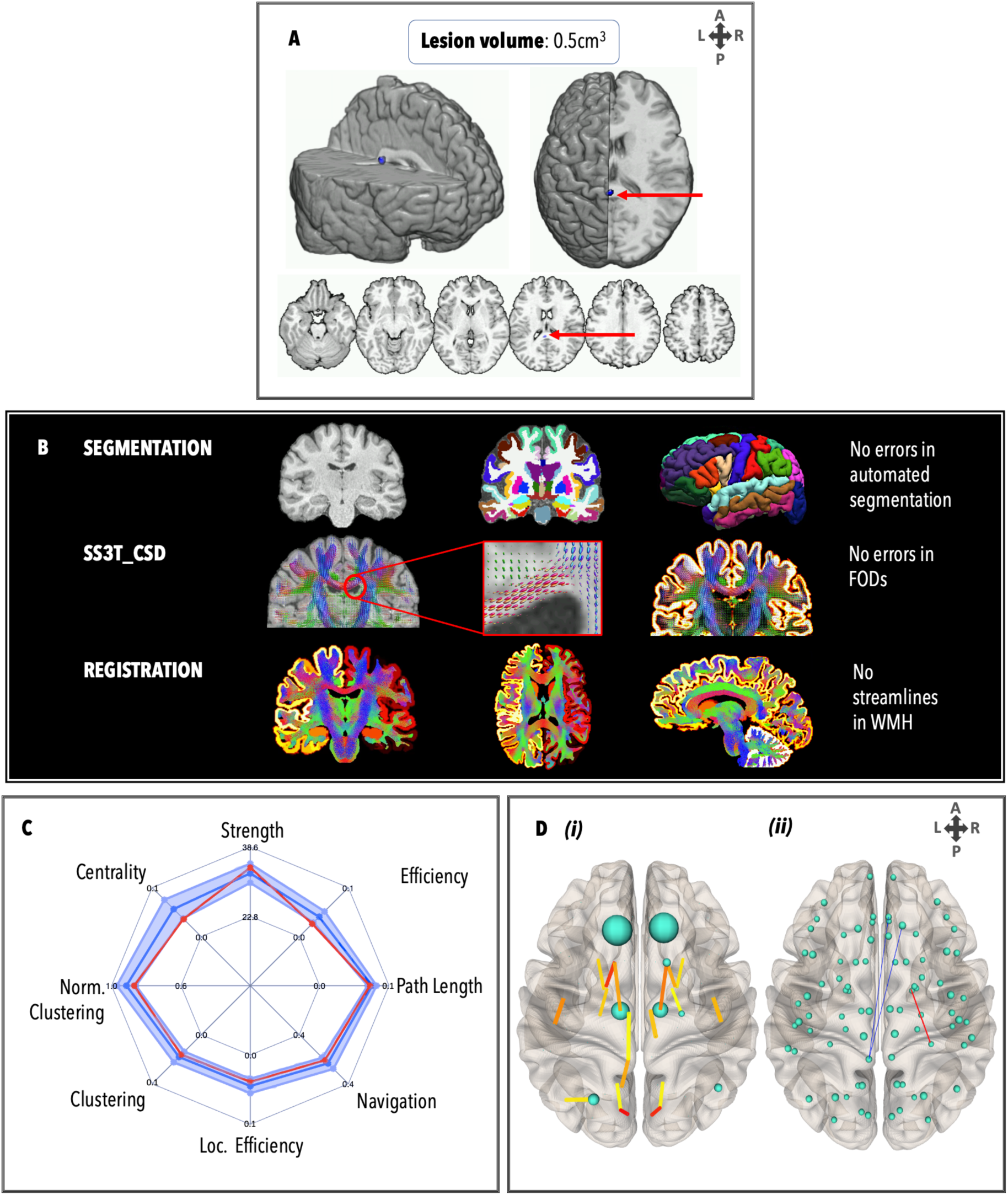
Personalised connectome profile for TBI6 including (A) lesion profile, (B) quality assessment, (C) radar plot showing the patient’s personalised connectome profile, and (D) regional analysis (blue=edges lower than healthy control average; red = edges stronger than healthy control average; and thicker edges=larger number of standard deviations away from healthy mean).

## Discussion

For the first time, we showcase an implementation of personalised connectomics in chronic moderate-to-severe TBI patients. In the following sections we discuss the defining characteristics of our single-subject profiles and explore ways in which our approach can contribute to improving existing methods of personalised structural connectome analyses in TBI patients.

### Single-subject network profiling observations

Our observations highlight a major caveat to approaches that attempt to identify a single graph metric that can be used as an adequate and parsimonious descriptor of structural network alterations in TBI patients^8^. In accordance with Lv and colleagues^13^, we observed that each TBI patient showed a unique *pattern* of graph metric alterations, regardless of lesion load. For example, while both TBI1 and TBI5 patients had small lesion loads, patient TBI1 had lower brain network integration and segregation measures, compared with non-significant deviation from the normal range for patient TBI5. By comparison, TBI3 and TBI4 both had much larger lesions, but patient TBI3’s brain network profile showed infra-normal integration and segregation measures, while patient TBI4’s brain network was supra-normal. Our results highlight the benefit of using a multivariate *profile* of graph metrics that reveal individual differences in brain network topology otherwise obscured by group-level analyses. Importantly, with the incorporation of individual edge and hub comparisons, the location of the lesion can be compared to edge deterioration in single patients.

### Improving methods for personalised connectomics

The present study introduces the structural connectivity aspect of a novel framework called STREAMLINES (Single-subjecT pRofiling of cognitivE impAirMents and structuraL braIN NEtwork metricS), which operates on state-of-the-art single-subject analyses of structural MRI scans that may provide clinicians with a novel user-friendly framework for leveraging graph metrics to benefit the individual patient. Advancing individual brain network profiling has the potential to inform neuroimaging-guided personalised rehabilitation programs by providing network-based summary statistics with prognostic capabilities^61-63^. More precisely, our approach can help to assess network alternations in TBI patients in the following three ways. First, regional connectome maps can be used as profiles of patients’ brain network topographies, thereby providing clinicians with time-efficient visual summaries of network disruption, asymmetry, hub alterations, and overall reductions in strength. Second, by comparing an individual patient to a healthy control reference group, we can observe portions of brain networks that are topologically altered but correspond to brain regions beyond the site of initial injuries. Finally, the GraphMe plots can be used longitudinally to map how the brain undergoes progressive secondary damage, recovery, and/or functional reorganisation over time^64,65^.

The current ‘best practice’ methods (lesion masking and manual editing^66^) are time-consuming and have low inter-rater reliability^67^. By contrast, use of the semi-automated lesion inpainting program VBG reduces the burden imposed by having to manually delineate lesions and avoids the exclusion of cases with large focal lesions that fail segmentation (e.g., from FreeSurfer)^19^. Furthermore, we observed that the SS3T-CSD model^46,68^ was suitable for constructing connectomes in the presence of lesions in all our TBI patients. SS3T-CSD removes the contributions from GM and cerebrospinal fluid (CSF) components to increase the specificity of FODs to the WM, while avoiding over-estimation into GM and CSF signal from the lesioned area^26^. Combined with anatomically constrained tractography^27^, streamlines are not generated in lesioned areas (e.g., see TBI1, Figure 3a panel C), and therefore anatomically disconnected regions do not have to be removed from connectivity matrices. This allowed us to calculate graph metrics from connectivity matrices of the same dimensions as those extracted from the healthy controls.

## Limitations

The implementation of personalised connectomics requires extensive validation and assessment of test-retest reliability. However, our study provides initial validation of this approach using six TBI patients and a small healthy control reference group (*N*=12)^12^. Normative analysis with a large (*N*>100), stratified sample of healthy individuals will allow stronger inferences to be made using techniques such as quartile regression^13, 11^. Personalised connectomics should also include a patient group as an additional reference cohort (*N*>100), to help clinicians understand how a patient is evolving with reference not only to healthy controls but also to patients with the same condition. Furthermore, our study only utilised T1 images for lesion identification; in the future, other structural imaging modalities such as fluid attenuated inversion recovery (FLAIR) and susceptibility weighted imaging (SWI) should also be used in accordance with best practice guidelines for lesion identification^69^. Despite multiple expert raters and use of an established procedure^22^, DAI grading remains subjective and requires independent confirmation of reliability. Hubs were defined according to previous work citing the use of betweenness centrality – however, other work has shown that finding ‘consensus’ between multiple centrality metrics may be more stable for hub definition^70^.Finally, cognitive outcomes associated with graph measures are still being evaluated – this progress will be essential for providing clinically informative personalised connectomes^23^ and is included as part of the STREAMLINES platform.

## Conclusions

Our results emphasize the translational potential for single-subject network analyses in the study of brain injury. Profiling individual patients based on their unique presentation provides insights into brain network disruption that are otherwise obscured by group-level approaches. The GraphMe profiling can provide clinicians with a novel user-friendly framework for leveraging graph metrics to benefit the individual patient by characterising network-level brain alterations with potential prognostic relevance. This study therefore enables us to progress towards a personalised medicine approach which, alongside group-based comparisons of patients against controls, is essential for translating connectomics to evidence-based clinical practice.

## Data Availability

All data produced in the present study are available upon reasonable request to the authors

## Acknowledgements

We would like to thank Dr. Alex Burmester for his help with the Hierarchical Drift Diffusion Modelling; and Michael Kean and the radiographers at the Royal Children’s Hospital.

## Authors’ contributions

*Imms, P:* Conceptualisation; Methodology; Software; Formal Analysis; Investigation; Data Curation; Writing – Original Draft; Writing – Review & Editing; Visualisation. *Clemente, A:* Investigation; Resources; Data Curation; Writing – Review & Editing. *Deutscher, E:* Formal Analysis; Writing – Review & Editing. *Ahmed Radwan:* Software; Resources. *Hamed Akhlaghi:* Resources; Data curation. *Beech, P:* Formal Analysis; Writing – Review & Editing. *Wilson, PH:* Conceptualisation; Resources; Writing – Review & Editing; Supervision; Funding acquisition. *Irimia, A:* Writing – Review & Editing. *Poudel, G:* Conceptualisation; Methodology; Software; Validation; Formal Analysis; Resources; Data Curation; Writing – Review & Editing; Visualisation; Supervision. *Domínguez D, JF:* Conceptualisation; Methodology; Software; Validation; Formal Analysis; Resources; Data Curation; Writing – Review & Editing; Supervision. *Caeyenberghs, K:* Conceptualisation; Methodology; Validation; Formal Analysis; Investigation; Resources; Writing – Original Draft; Writing – Review & Editing; Supervision; Project Administration; Funding Acquisition.

