## Supplementary Materials for "Personalised structural connectomics for moderate-to-severe traumatic brain injury"

### Supplementary Material 1

***Edge reconstruction***

To estimate the white matter fibre orientation distributions (FODs) in each voxel, single-shell 3-tissue constrained spherical deconvolution (SS3T-CSD) was performed[25]. SS3T-CSD preserves the angular information of the GM- and CSF-like signal, removing contributions from these components to increase the specificity of the WM FODs, while avoiding overestimation into GM and CSF signal from the lesioned area[26]. Whole-brain, anatomically-constrained tractography was performed using a 5 tissue-type segmentation of the T1 images in dMRI space to create the relevant masks for tractography[27]. Twenty-two million streamlines were generated to keep connectome variability low enough for SIFT2 to be relatively stable[28]. The SIFT2 algorithm was applied to match the density of the reconstructed streamlines to that of the underlying white matter structures[28-30]. A proportionality coefficient μ was also calculated for each participant to be later applied to the connectome edge weights to ensure these are proportional to the apparent fibre density.

***Node construction***

Anatomical images were parcellated using FreeSurfer's *recon-all* function (v6.0; http://surfer.nmr.mgh.harvard.edu/), as described in previous publications[32]. On this surface model the automated cortical and subcortical parcellation of 84 regions was generated using the Desikan-Killiany atlas[31]. Quality control was performed by inspecting output of the FreeSurfer pipeline at each stage using stringent ENIGMA guidelines (http://enigma.usc.edu/). Two patients (TBI3 and TBI4) did not pass the quality checks, due to significant segmentation failures in the presence of pathology, and were analysed utilising the new virtual brain grafting (VBG v0.37) image processing pipeline to improve segmentation[19]. Lesions are filled with healthy tissue from synthetic 'donor brain' images – either leveraging tissue from the native non-lesioned hemisphere for unilateral lesions, or a healthy synthetic donor brain for bilateral lesions. The resulting lesion free patient image is then provided as input to the FreeSurfer *recon-all* pipeline, thereby enabling improved segmentation in the absence of structural pathology.

**Graph Theoretical Analysis**

Connectivity matrices were generated using edge weights from SIFT2 and brain regions from FreeSurfer and VBG. Area size normalisation occurred by scaling to the inverse volumes of the nodes they connect[71]. Network architecture was quantified in terms of *strength*, *global efficiency, characteristic path length*, *navigation efficiency*, *local efficiency, clustering coefficient, normalised clustering coefficient,* and *betweenness centrality* (Table 2), using the Brain Connectivity Toolbox[16]. Graph normalisation occurred by 1) normalising edge weights between 0 and 1, and 2) weight-to-length remapping using -log transformation (for global efficiency, characteristic path length, local efficiency, and navigation efficiency only). Graph metrics were calculated for each TBI patient individually and for the group connectivity matrix of the healthy control subjects.

**Supplementary Material 2**

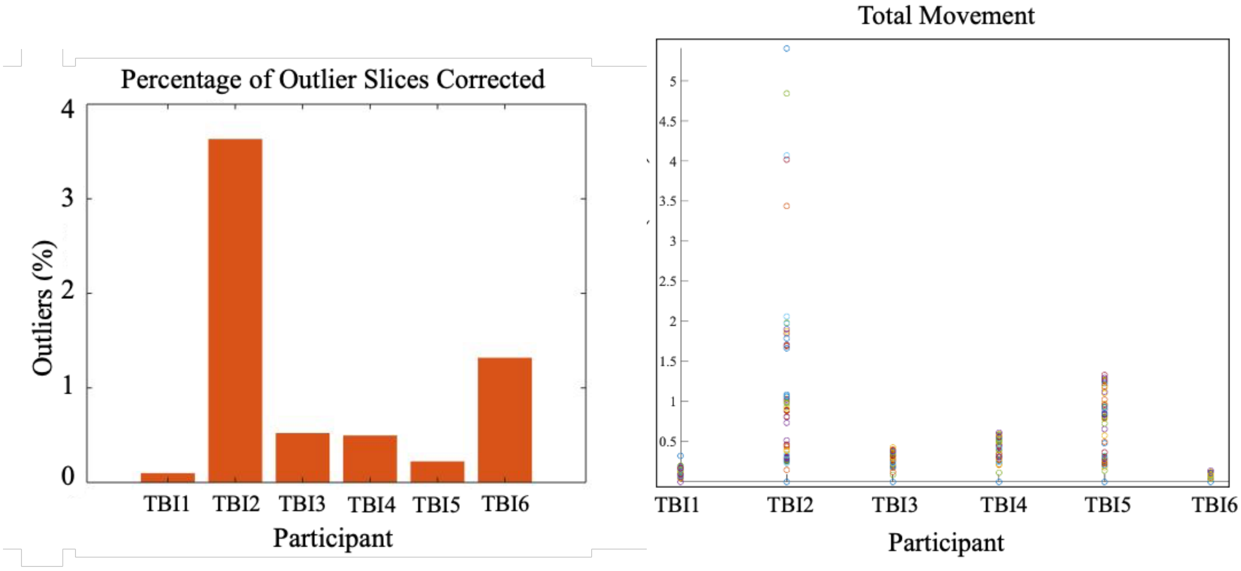


**Supp Fig 2.** Head motion summary for six TBI patients. Outlier slices are corrected by removing slices and replacing with corrected slices; Data points in the Total Movement plot represent the root mean square (*rms*) deviation from centre of mass for each volume ($n$=67). Slices that had greater movement than two standard deviations from the average were replaced automatically by FSL's outlier correction[72]. After outlier correction, motion values were below the voxel size of image acquisition (before upsampling) for each patient except TBI2 (*rms* = 5.41 mm); this patient was excluded from subsequent diffusion imaging analyses.

**Supplementary Material 3**

**TBI2**

TBI2 was excluded from further diffusion MRI analysis due to head motion greater than the voxel size of image acquisition. TBI2 demonstrated extensive bilateral frontal, and right parietal and temporal lesions (load = 163cm$^3$), as well as focal hypointensities in the left thalamus and body and genu of the corpus callosum, and a DAI grade of 2. Prior to VBG, 22 nodes failed the quality assessment. VBG improved segmentation in 15 nodes. The remaining 7 nodes are located predominantly in lesioned areas. Constrained spherical deconvolution based on the single-shell 3 tissue FODs was not generated at the site of the lesions (see red arrow), and registration between VBG repaired nodes and streamlines show that streamlines were not assigned to lesioned nodes.

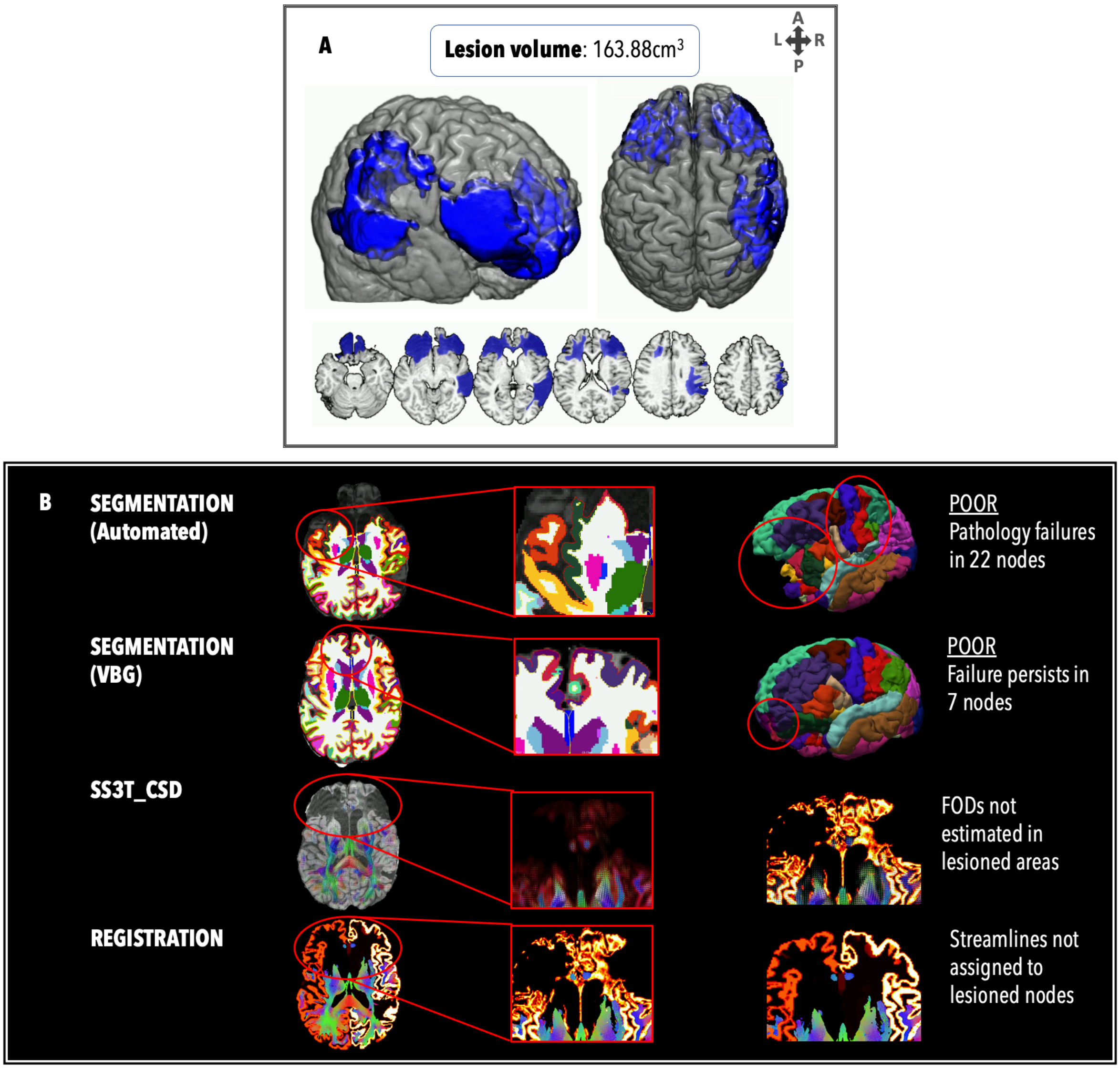


**Supp Fig 3.** Personalised connectome profile for TBI2 including (A) lesion profile; (B) quality assessment; (C) radar plot showing the patient's personalised connectome profile (red indicates patient's scores, dark blue indicates healthy control average and the 95% Cis are represented by the blue shade); and (D) regional analysis (blue: edges lower than the healthy control average; red: edges stronger than the healthy control average; thicker edges: more standard deviations away from the healthy mean).
